# Urinary Sodium and Incident Apparent Treatment-Resistant Hypertension among African American adults: The JHS

**DOI:** 10.1101/2024.10.23.24316022

**Authors:** Olutobi A. Sanuade, Daniel K Addo, Justin D. Smith, Allison J. Carroll, Daichi Shimbo, Sameera Talegawkar, Katherine L. Tucker, Joshua A. Jacobs, Catherine G. Derington, Adam P. Bress

## Abstract

Hypertension is a leading cause of cardiovascular disease and disproportionately affects African American (AA) adults. Apparent treatment-resistant hypertension (aTRH) is highly prevalent in this population. Sodium intake is associated with blood pressure (BP) levels, yet the relationship between sodium and aTRH in AA adults remains unclear. This study examined the association between 24-hour urinary sodium excretion and incident aTRH among AA adults with hypertension, using data from the Jackson Heart Study (JHS). The JHS included 5,306 self-identified AA adults from Jackson, Mississippi, with data collected from 2000 to 2013. This analysis included 452 participants with baseline hypertension and complete urinary excretion and medication data. Sodium excretion was categorized into quartiles based on their urinary excretion: Q1 (253 to 2530 mg/day), Q2 (2553 to 3657 mg/day), Q3 (3680 to 4692 mg/day), and Q4 (4715 to 9775 mg/day). aTRH was defined as uncontrolled BP while taking ≥3 antihypertensive medications. A semi-parametric proportional hazards model was used to determine the association between sodium excretion and aTRH, adjusting for confounders. Participants in the current analyses were 63 years old on average and 27.7% men. Over a median follow-up of 7.5 years, 123 participants (27.2%) developed aTRH. Participants in Q3 and Q4 of sodium excretion showed higher incidence of aTRH, though fully adjusted hazard ratios were not statistically significant [HRs (95% confidence intervals [CIs]): [Q2=0.71 (0.34, 1.46), Q3=1.02 (0.50, 2.06), Q4=0.95 (0.46, 2.00); P=0.166). There was no statistically significant association between urinary sodium and incident aTRH among AA adults with hypertension.

## Background

Almost 1 in 3 American adults has high blood pressure (BP),^1^ with less than half achieving adequate BP control, increasing their risk for heart disease and stroke. Hypertension contributes to 410,000 deaths annually and disproportionately affects minority populations, particularly African American (AA) adults, with significant financial and health impacts.^2^ AA adults have higher rates of hypertension compared to white Americans, contributing to a 5.5-year reduction in life expectancy for AA adults.^3^

Reducing sodium intake is an effective strategy to lower BP and improve cardiovascular health.^4–6^ The American Heart Association (AHA) recommends sodium intake of <2300 mg/day. However, most Americans consume an average of 3,400 mg/day,^7^ with 86% of adults with hypertension exceeding the recommended limit.^8^ AA adults consume more sodium than other racial or ethnic populations in the US due to greater consumption of processed foods, restaurant meals, salt added to home-cooked meals,^9^ along with systemic barriers like poverty.^10^ Urinary sodium excretion is a reliable measure of dietary sodium intake,^11^ making it a robust measure for assessing sodium consumption.^12^

Apparent treatment resistant hypertension (aTRH), defined as BP above target despite use of ≥3 classes of antihypertensive medication or ≥4 classes regardless of BP level, is a common clinical problem^13^ and associated with increased cardiovascular risk.^14^ AA adults are more likely to have resistant hypertension due to genetic predisposition to salt and water retention than white adults.^15^

Current literature lacks robust evidence on the association between sodium intake and aTRH specifically in AA adults, who have hypertension. Studies demonstrating that reduced sodium intake significantly lowers BP in patients with resistant hypertension^4,5^ have limited external validity due to small sample sizes. Addressing these gaps is important for developing dietary and clinical interventions for this high-risk population. This study examined the association of sodium intake with incident aTRH among AA adults with hypertension from the Jackson Heart Study (JHS). We hypothesize a positive linear association between sodium intake, as measured by urinary sodium, and incident aTRH in this population.

## METHODS

### Study Population

The JHS is a prospective community-based observational study of risk factors for cardiovascular disease in AA adults. The JHS recruited 5,306 adults aged 21-94 years, residing in the tri-county area of the Jackson, Mississippi, USA, metropolitan area. Participants were examined at baseline – Visit 1 (2000–2004) and two additional examinations: Visit 2 (2005–2008) and Visit 3 (2009–2013). Further details of the JHS study design, recruitment, and data collection were published previously.^16–18^ The study was approved by the Institutional Review Boards of Jackson State University, Tougaloo College, and the University of Mississippi Medical Center in Jackson. All participants provided written informed consent. This article was reviewed by the JHS Publications and Presentations Committee and has been approved for publication.

### Inclusion and Exclusion Criteria

The JHS cohort included 5,306 participants at Visit 1. We excluded 4,854 participants missing 24-hour urinary sodium data at Visit 1 (n=4,280), BP medication data (n=17), or hypertension diagnosis (n=371) and those with prevalent aTRH at baseline (n=186) (**Figure 1**). After applying these exclusion criteria, we included 452 JHS participants with hypertension (defined as SBP ≥ 130 mm Hg or DBP ≥ 80 mm Hg, according to the 2017 ACC/AHA BP guidelines hypertension^14^) in the present analyses.

**Figure 1.**
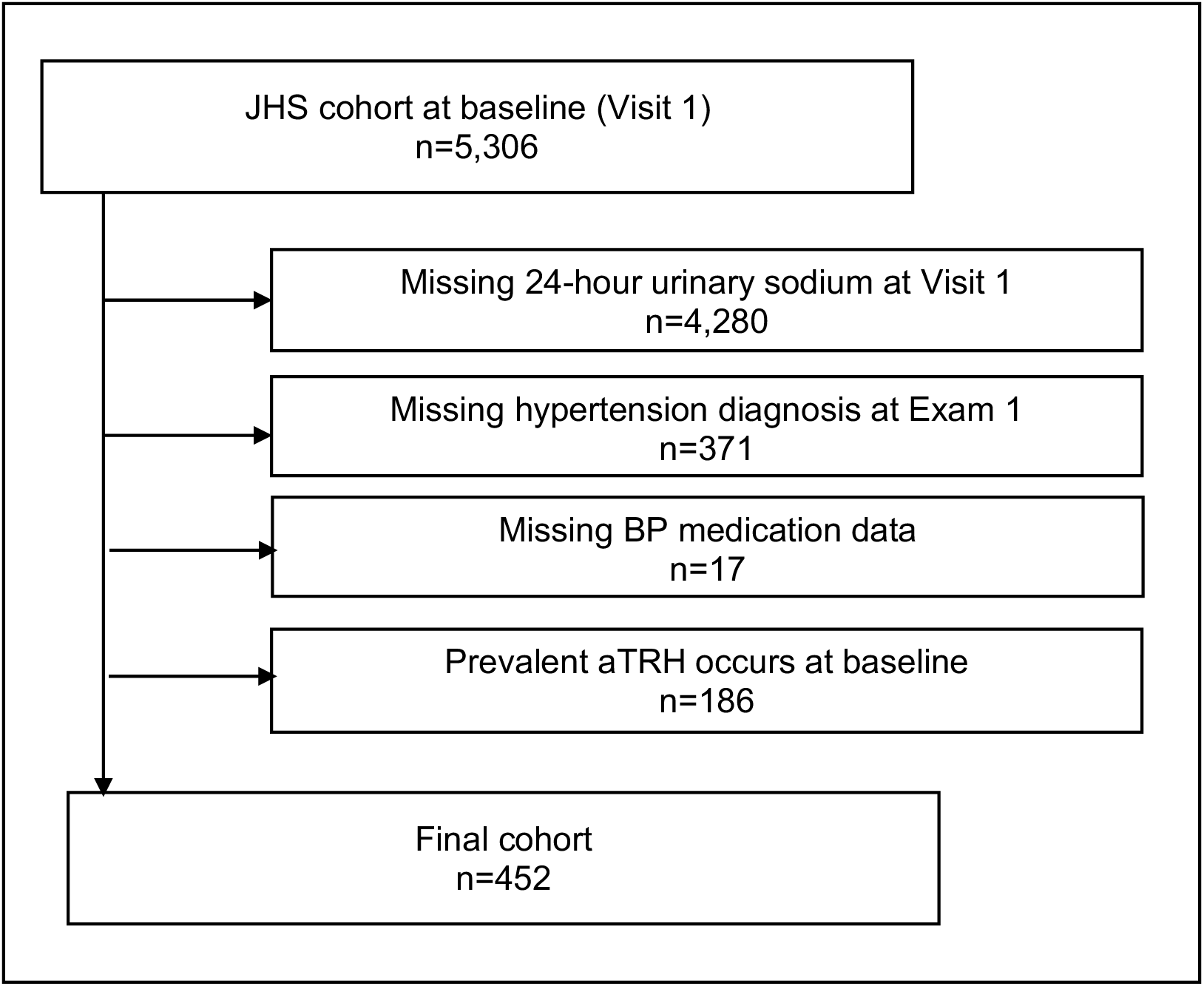
Flowchart showing the eligibility criteria applied to participants in the Jackson Heart Study (JHS) for the analysis of incident aTRH.

### Data collection

JHS data were collected in two phases: an in-home interview followed by a clinical examination at the JHS clinic. During the in-home interview, participants provided information on their demographics (i.e., age, sex, education, income, and insurance status), lifestyle factors (smoking status, alcohol consumption, and physical activity), medical history (i.e., coronary heart disease history, stroke history, myocardial infarction history, chronic kidney disease history, and diabetes history) and medication use (i.e., statin use, and antihypertensive medication use– angiotensin-converting enzyme inhibitors–ACEI, angiotensin II receptor blockers–ARB, beta-blockers, calcium channel blockers–CCB, diuretics, and other hypertensive medication classes).^18^ The JHS physical activity instrument was also administered to assess participants’ physical activity levels as poor, intermediate, or ideal/recommended.^19,20^

During the clinic visit, medication adherence was assessed by reviewing the participants’ pill bottles to record the medications taken in the two weeks before the visit. A standardized protocol was used to conduct physical assessments and collect biological samples. BP, height, and weight were measured, and blood and urine samples were obtained. BP was based on the average of two seated measurements, taken with an appropriate cuff size and a standard Hawksley random zero sphygmomanometer (Hawksley and Sons Limited). Participants rested for five minutes before the first measurement, with a 30-second interval between measurements. The raw BP data obtained from the random zero sphygmomanometers were subsequently calibrated using robust regression models.^21^ Total cholesterol, low-density lipoprotein (LDL), and high-density lipoprotein (HDL) levels were measured from fasting blood samples using standardized enzymatic methods. Body mass index (BMI) was calculated as weight (kg)/height (m)^2^. Fasting serum glucose was measured using the glucose oxidase method on a Vitros 950 or 250 analyzer (Ortho Clinical Diagnostics). Glycated hemoglobin was assessed using a Tosoh high-performance liquid chromatography system (Tosoh Corporation).

Further, urine samples were analyzed to measure sodium intake and albumin levels using the Dade Behring BN II nephelometer (Siemens), based on the specimens collected during the baseline study visit.^22^ Participants were provided with instructions and a urine collection kit to complete a 24-hour urine collection. JHS staff verbally ensured that participants understood the collection procedures, and participants collected all urine over a 24-hour period, with collection jars labeled with participant ID, and collection times. Serum and urine creatinine concentrations were measured using a multi-point enzymatic spectrophotometric assay on a Vitros 950 Ortho Clinical Diagnostics analyzer. For analysis purposes, the creatinine values were biochemically calibrated to the Cleveland Clinic–equivalent Minnesota Beckman CX3 assay.^22,23^ Urinary albumin-to-creatinine ratio (ACR) was generated by dividing the albumin levels by the creatinine values. Estimated glomerular filtration rate (eGFR) was calculated via the Chronic Kidney Disease Epidemiology Collaboration (CKD-EPI) equation.^24^ C-reactive protein (CRP) was assessed using the immunoturbidimetric CRP-Latex assay on a Hitachi 911 analyzer, utilizing high-sensitivity C-reactive protein (hs-CRP) measurements that can detect smaller increases in CRP compared to standard tests.^25^

### Exposure

The main exposure was 24-hour urine sodium excretion, expressed as milligrams (mg) at Visit 1. Participants were stratified into quartiles based on their urinary sodium levels: Q1 (n=113, 253 – 2530 mg/day); Q2 (n=113, 2553 – 3657 mg/day); Q3 (n=113, 3680 – 4692 mg/day), and; Q4 (n=113, 4715 – 9775 mg/day).

### Outcome

The outcome variable was incident aTRH at visits 2 or 3. Participants with uncontrolled BP (i.e., SBP ≥ 130 mm Hg or DBP ≥ 80 mm Hg, according to the 2017 ACC/AHA BP guidelines and the 2018 AHA Scientific Statement on resistant hypertension^14^) while taking three classes of antihypertensive medication or controlled BP (i.e., SBP < 130 mm Hg and DBP < 80 mm Hg) while taking four or more classes of antihypertensive medication, were classified with aTRH.^13,14,22^

### Covariates

Covariates included demographic variables, lifestyle factors, antihypertensive medication use, medication adherence, BMI, fasting cholesterol (total, low-density lipoprotein–LDL and high-density lipoprotein–HDL), urinary albumin-to-creatinine ratio (ACR), and C-reactive protein (CRP).

### Statistical analysis

Baseline characteristics across quartiles of urinary sodium were examined. Normally distributed continuous data were presented as mean ± standard deviation, while non-normally distributed continuous data were expressed as median (Q1, Q3). Categorical variables were presented as percentages. We used histograms to display the baseline distribution of 24-h urinary sodium levels (**Supplemental Figure 1**). Missingness in baseline characteristics are presented in **Supplemental Table 1**.

To account for missingness in the baseline covariates, we used multiple imputation via chained equations with guidelines relevant for interval-censored data.^26,27^ Briefly, we included in our imputation model all baseline covariates, the exposure, aTRH event indicator, and the cumulative baseline hazard. We had at least as many imputed datasets as the percentage of individuals with any missing data (approximately 24%). Rubin’s rules were adopted for combining estimates and standard errors across the imputed datasets.^28^

We examined the association between 24-h urinary sodium excretion and incident aTRH using a semi-parametric proportional hazards regression model for interval-censored data,^29^ adjusting for potential confounders. In Model 1, we adjusted for age and sex. For Model 2, additional adjustments were made for smoking status, alcohol consumption, physical activity, BMI, diabetes, history of coronary heart disease, history of stroke, heart failure, chronic kidney disease, estimated glomerular filtration rate, low– and high-density lipoproteins, urinary albumin-to-creatinine ratio, medication adherence, and an indicator for each antihypertensive medication class. In Model 3, we made additional adjustments for income, education and insurance status.

Hazard ratios (HRs) for incident aTRH were estimated using quartiles of urinary sodium, with the lowest quartile (Q1) as the reference. We visualized cumulative incidence curves for incident aTRH by quartiles of urinary sodium using Turnbull’s estimator.^30^ Participants who did not develop aTRH were right censored at their last follow-up visit, while those who developed aTRH were left or interval-censored, as appropriate. Assuming a proportional hazards model, we graphed HRs for incident aTRH associated with urinary sodium using restricted quadratic splines with a single knot specified at the median value. Analyses were conducted using R version 4.3.2.^31^ We used the *mice* and *icenReg* packages available in R to run multiple imputation and fit all models appropriate for interval-censored data respectively.^32,33^

## RESULTS

### Participant Characteristics

The baseline characteristics of the 452 included participants are presented in **Table 1** (median age 63.0 years; 27.7% men). Participants were stratified into four groups based on their 24-h urinary sodium excretion levels: Q1 (253 to 2530 mg/day), Q2 (2553 to 3657 mg/day), Q3 (3680 to 4692 mg/day), and Q4 (4715 to 9775 mg/day). The overall median 24-h urinary sodium excretion was 3,669 (2,547, 4,698) mg/day and the distribution is presented in **Supplemental Figure 1**.

**Table 1.**
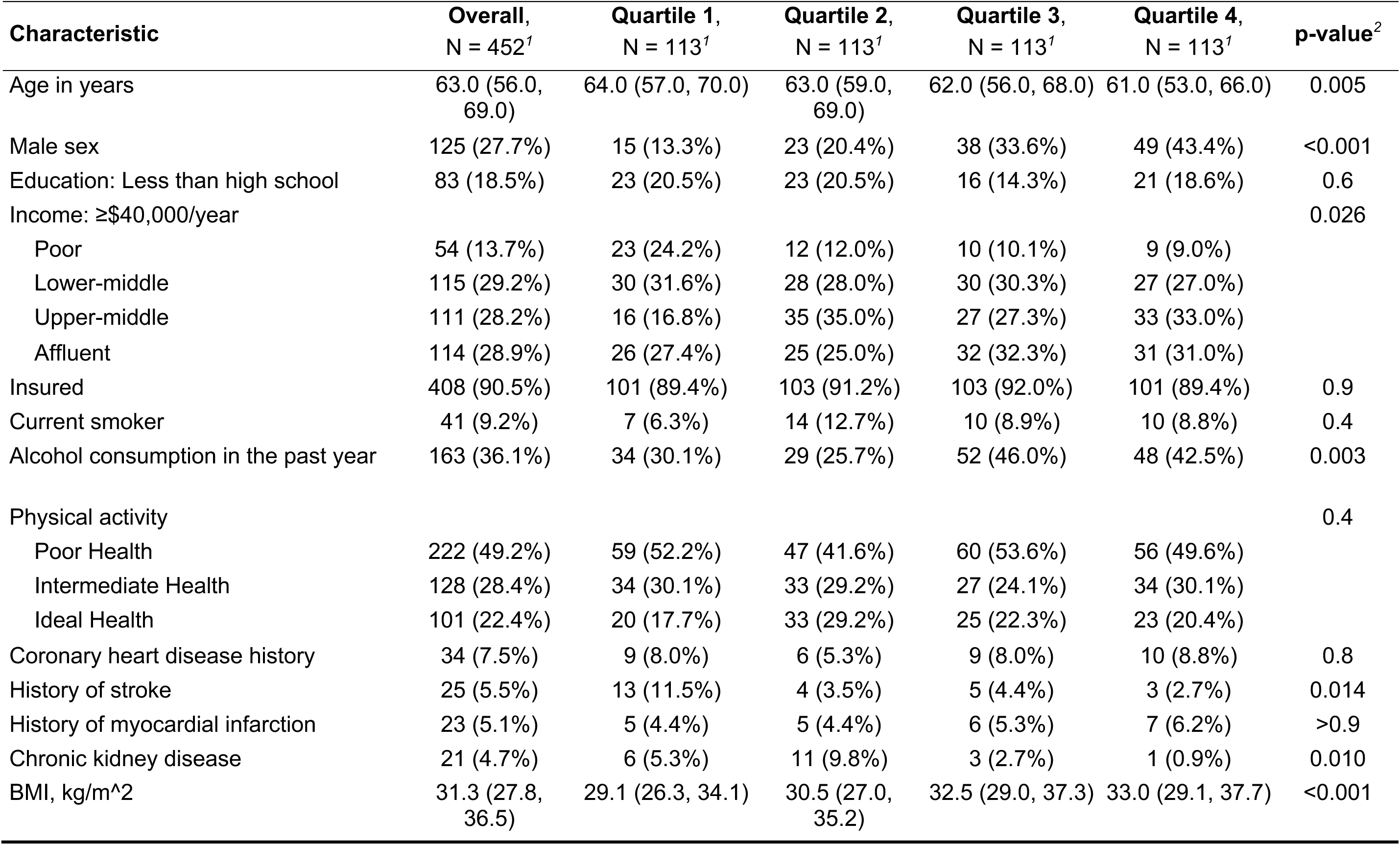
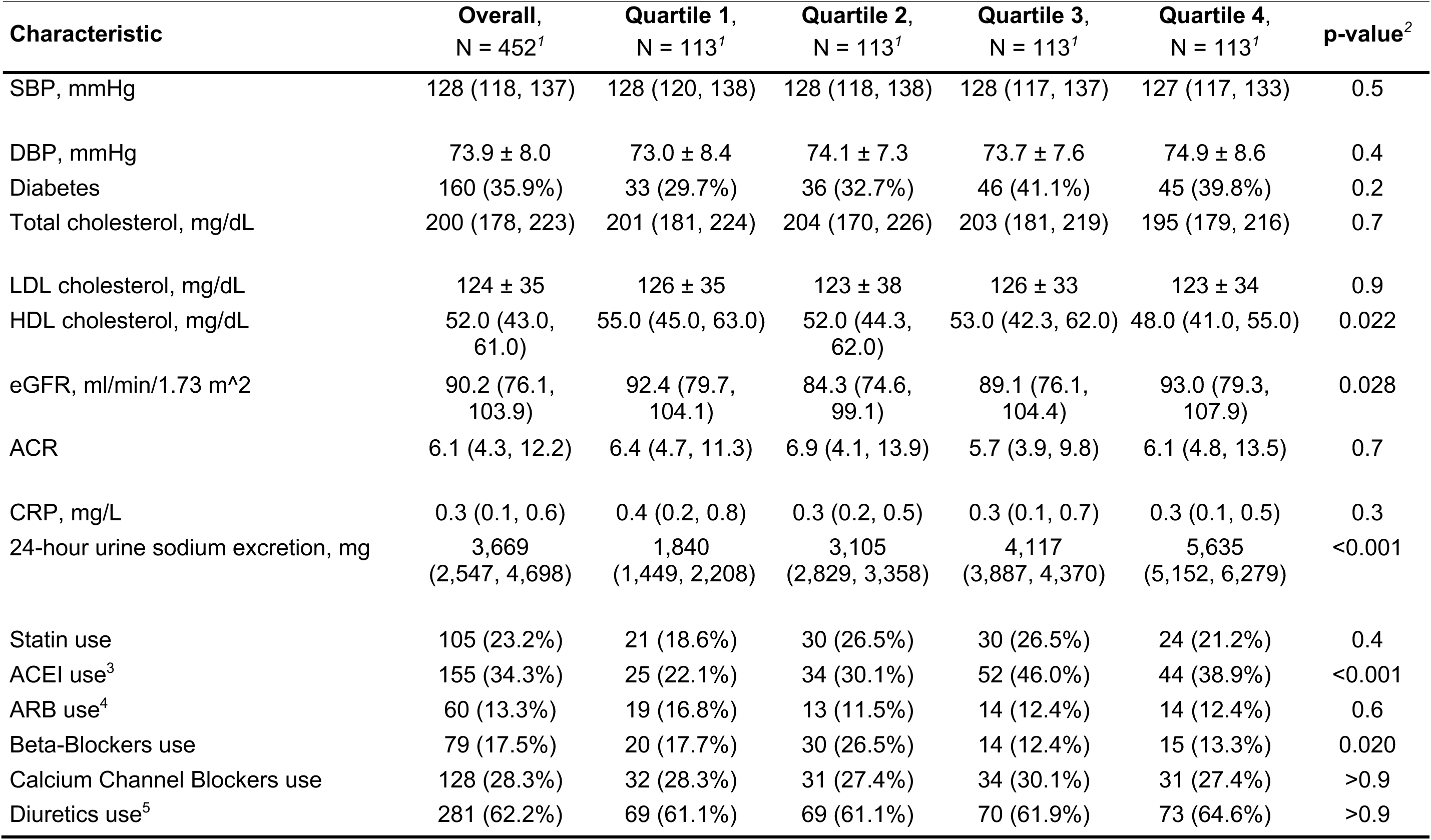
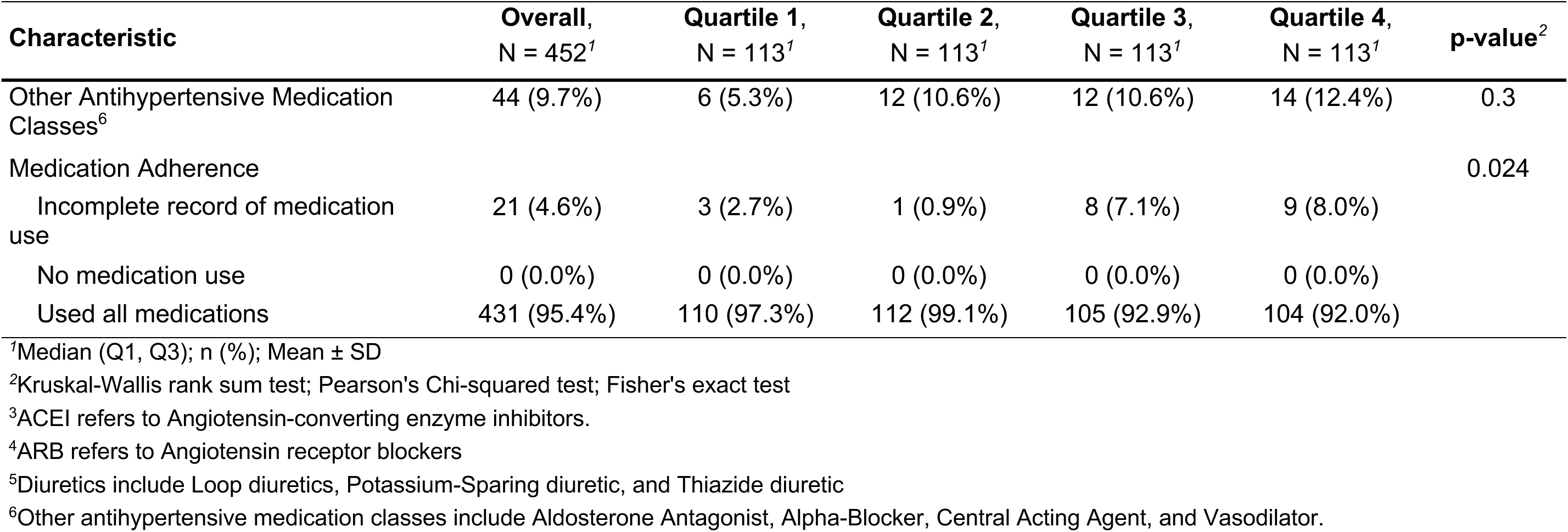
Baseline characteristics of JHS participants enrolled in 2000–2004 and included in the analysis of incident aTRH overall by quartiles of urinary sodium.

| Characteristic | Overall,<br>N = 452<sup>1</sup> | Quartile 1,<br>N = 113<sup>1</sup> | Quartile 2,<br>N = 113<sup>1</sup> | Quartile 3,<br>N = 113<sup>1</sup> | Quartile 4,<br>N = 113<sup>1</sup> | p-value<sup>2</sup> |
| --- | --- | --- | --- | --- | --- | --- |
| Age in years | 63.0 (56.0, 69.0) | 64.0 (57.0, 70.0) | 63.0 (59.0, 69.0) | 62.0 (56.0, 68.0) | 61.0 (53.0, 66.0) | 0.005 |
| Male sex | 125 (27.7%) | 15 (13.3%) | 23 (20.4%) | 38 (33.6%) | 49 (43.4%) | <0.001 |
| Education: Less than high school | 83 (18.5%) | 23 (20.5%) | 23 (20.5%) | 16 (14.3%) | 21 (18.6%) | 0.6 |
| Income: ≥\$40,000/year | | | | | | 0.026 |
| Poor | 54 (13.7%) | 23 (24.2%) | 12 (12.0%) | 10 (10.1%) | 9 (9.0%) |  |
| Lower-middle | 115 (29.2%) | 30 (31.6%) | 28 (28.0%) | 30 (30.3%) | 27 (27.0%) |  |
| Upper-middle | 111 (28.2%) | 16 (16.8%) | 35 (35.0%) | 27 (27.3%) | 33 (33.0%) |  |
| Affluent | 114 (28.9%) | 26 (27.4%) | 25 (25.0%) | 32 (32.3%) | 31 (31.0%) |  |
| Insured | 408 (90.5%) | 101 (89.4%) | 103 (91.2%) | 103 (92.0%) | 101 (89.4%) | 0.9 |
| Current smoker | 41 (9.2%) | 7 (6.3%) | 14 (12.7%) | 10 (8.9%) | 10 (8.8%) | 0.4 |
| Alcohol consumption in the past year | 163 (36.1%) | 34 (30.1%) | 29 (25.7%) | 52 (46.0%) | 48 (42.5%) | 0.003 |
| Physical activity |  |  |  |  |  | 0.4 |
| Poor Health | 222 (49.2%) | 59 (52.2%) | 47 (41.6%) | 60 (53.6%) | 56 (49.6%) |  |
| Intermediate Health | 128 (28.4%) | 34 (30.1%) | 33 (29.2%) | 27 (24.1%) | 34 (30.1%) |  |
| Ideal Health | 101 (22.4%) | 20 (17.7%) | 33 (29.2%) | 25 (22.3%) | 23 (20.4%) |  |
| Coronary heart disease history | 34 (7.5%) | 9 (8.0%) | 6 (5.3%) | 9 (8.0%) | 10 (8.8%) | 0.8 |
| History of stroke | 25 (5.5%) | 13 (11.5%) | 4 (3.5%) | 5 (4.4%) | 3 (2.7%) | 0.014 |
| History of myocardial infarction | 23 (5.1%) | 5 (4.4%) | 5 (4.4%) | 6 (5.3%) | 7 (6.2%) | >0.9 |
| Chronic kidney disease | 21 (4.7%) | 6 (5.3%) | 11 (9.8%) | 3 (2.7%) | 1 (0.9%) | 0.010 |
| BMI, kg/m<sup>2</sup> | 31.3 (27.8, 36.5) | 29.1 (26.3, 34.1) | 30.5 (27.0, 35.2) | 32.5 (29.0, 37.3) | 33.0 (29.1, 37.7) | <0.001 |

| Characteristic | Overall,<br>N = 452<sup>1</sup> | Quartile 1,<br>N = 113<sup>1</sup> | Quartile 2,<br>N = 113<sup>1</sup> | Quartile 3,<br>N = 113<sup>1</sup> | Quartile 4,<br>N = 113<sup>1</sup> | p-value<sup>2</sup> |
| --- | --- | --- | --- | --- | --- | --- |
| SBP, mmHg | 128 (118, 137) | 128 (120, 138) | 128 (118, 138) | 128 (117, 137) | 127 (117, 133) | 0.5 |
| DBP, mmHg | 73.9 ± 8.0 | 73.0 ± 8.4 | 74.1 ± 7.3 | 73.7 ± 7.6 | 74.9 ± 8.6 | 0.4 |
| Diabetes | 160 (35.9%) | 33 (29.7%) | 36 (32.7%) | 46 (41.1%) | 45 (39.8%) | 0.2 |
| Total cholesterol, mg/dL | 200 (178, 223) | 201 (181, 224) | 204 (170, 226) | 203 (181, 219) | 195 (179, 216) | 0.7 |
| LDL cholesterol, mg/dL | 124 ± 35 | 126 ± 35 | 123 ± 38 | 126 ± 33 | 123 ± 34 | 0.9 |
| HDL cholesterol, mg/dL | 52.0 (43.0, 61.0) | 55.0 (45.0, 63.0) | 52.0 (44.3, 62.0) | 53.0 (42.3, 62.0) | 48.0 (41.0, 55.0) | 0.022 |
| eGFR, ml/min/1.73 m<sup>2</sup> | 90.2 (76.1, 103.9) | 92.4 (79.7, 104.1) | 84.3 (74.6, 99.1) | 89.1 (76.1, 104.4) | 93.0 (79.3, 107.9) | 0.028 |
| ACR | 6.1 (4.3, 12.2) | 6.4 (4.7, 11.3) | 6.9 (4.1, 13.9) | 5.7 (3.9, 9.8) | 6.1 (4.8, 13.5) | 0.7 |
| CRP, mg/L | 0.3 (0.1, 0.6) | 0.4 (0.2, 0.8) | 0.3 (0.2, 0.5) | 0.3 (0.1, 0.7) | 0.3 (0.1, 0.5) | 0.3 |
| 24-hour urine sodium excretion, mg | 3,669<br>(2,547, 4,698) | 1,840<br>(1,449, 2,208) | 3,105<br>(2,829, 3,358) | 4,117<br>(3,887, 4,370) | 5,635<br>(5,152, 6,279) | <0.001 |
| Statin use | 105 (23.2%) | 21 (18.6%) | 30 (26.5%) | 30 (26.5%) | 24 (21.2%) | 0.4 |
| ACEI use<sup>3</sup> | 155 (34.3%) | 25 (22.1%) | 34 (30.1%) | 52 (46.0%) | 44 (38.9%) | <0.001 |
| ARB use<sup>4</sup> | 60 (13.3%) | 19 (16.8%) | 13 (11.5%) | 14 (12.4%) | 14 (12.4%) | 0.6 |
| Beta-Blockers use | 79 (17.5%) | 20 (17.7%) | 30 (26.5%) | 14 (12.4%) | 15 (13.3%) | 0.020 |
| Calcium Channel Blockers use | 128 (28.3%) | 32 (28.3%) | 31 (27.4%) | 34 (30.1%) | 31 (27.4%) | >0.9 |
| Diuretics use<sup>5</sup> | 281 (62.2%) | 69 (61.1%) | 69 (61.1%) | 70 (61.9%) | 73 (64.6%) | >0.9 |

| Characteristic | Overall,<br>N = 452<sup>1</sup> | Quartile 1,<br>N = 113<sup>1</sup> | Quartile 2,<br>N = 113<sup>1</sup> | Quartile 3,<br>N = 113<sup>1</sup> | Quartile 4,<br>N = 113<sup>1</sup> | p-value<sup>2</sup> |
| --- | --- | --- | --- | --- | --- | --- |
| Other Antihypertensive Medication Classes<sup>6</sup> | 44 (9.7%) | 6 (5.3%) | 12 (10.6%) | 12 (10.6%) | 14 (12.4%) | 0.3 |
| Medication Adherence |  |  |  |  |  | 0.024 |
| Incomplete record of medication use | 21 (4.6%) | 3 (2.7%) | 1 (0.9%) | 8 (7.1%) | 9 (8.0%) |  |
| No medication use | 0 (0.0%) | 0 (0.0%) | 0 (0.0%) | 0 (0.0%) | 0 (0.0%) |  |
| Used all medications | 431 (95.4%) | 110 (97.3%) | 112 (99.1%) | 105 (92.9%) | 104 (92.0%) |  |

<sup>1</sup>Median (Q1, Q3); n (%); Mean ± SD
<sup>2</sup>Kruskal-Wallis rank sum test; Pearson's Chi-squared test; Fisher's exact test
<sup>3</sup>ACEI refers to Angiotensin-converting enzyme inhibitors.
<sup>4</sup>ARB refers to Angiotensin receptor blockers
<sup>5</sup>Diuretics include Loop diuretics, Potassium-Sparing diuretic, and Thiazide diuretic
<sup>6</sup>Other antihypertensive medication classes include Aldosterone Antagonist, Alpha-Blocker, Central Acting Agent, and Vasodilator.

Compared to those with lower urinary sodium levels, participants with higher urinary sodium levels were more likely to be younger, female, have higher income, consume alcohol, and use any antihypertensive medication class. Additionally, higher urinary sodium was associated with higher BMI, higher eGFR levels, or lower HDL cholesterol levels. Conversely, higher urinary sodium levels were less common among participants with a history of stroke, chronic kidney disease and those using ACEIs and beta-blockers.

### Association between Urinary sodium and incident aTRH

**Table 2** presents the incidence rates and HRs for aTRH, by quartile. Over a median follow-up of ∼7.5 years, 123 participants (27.2%) developed aTRH. Q3 and Q4 showed the highest incident aTRH, each with 33 cases (29.2%), while Q1 had the lowest incidence, with 29 cases (25.7%). The cumulative incidence curve for incident aTRH by quartiles of 24-h urinary sodium excretion (**Figure 2**) shows that individuals with higher urinary sodium levels (Q3 and Q4) had higher risk of developing aTRH over time compared to those with lower sodium levels.

**Figure 2.**
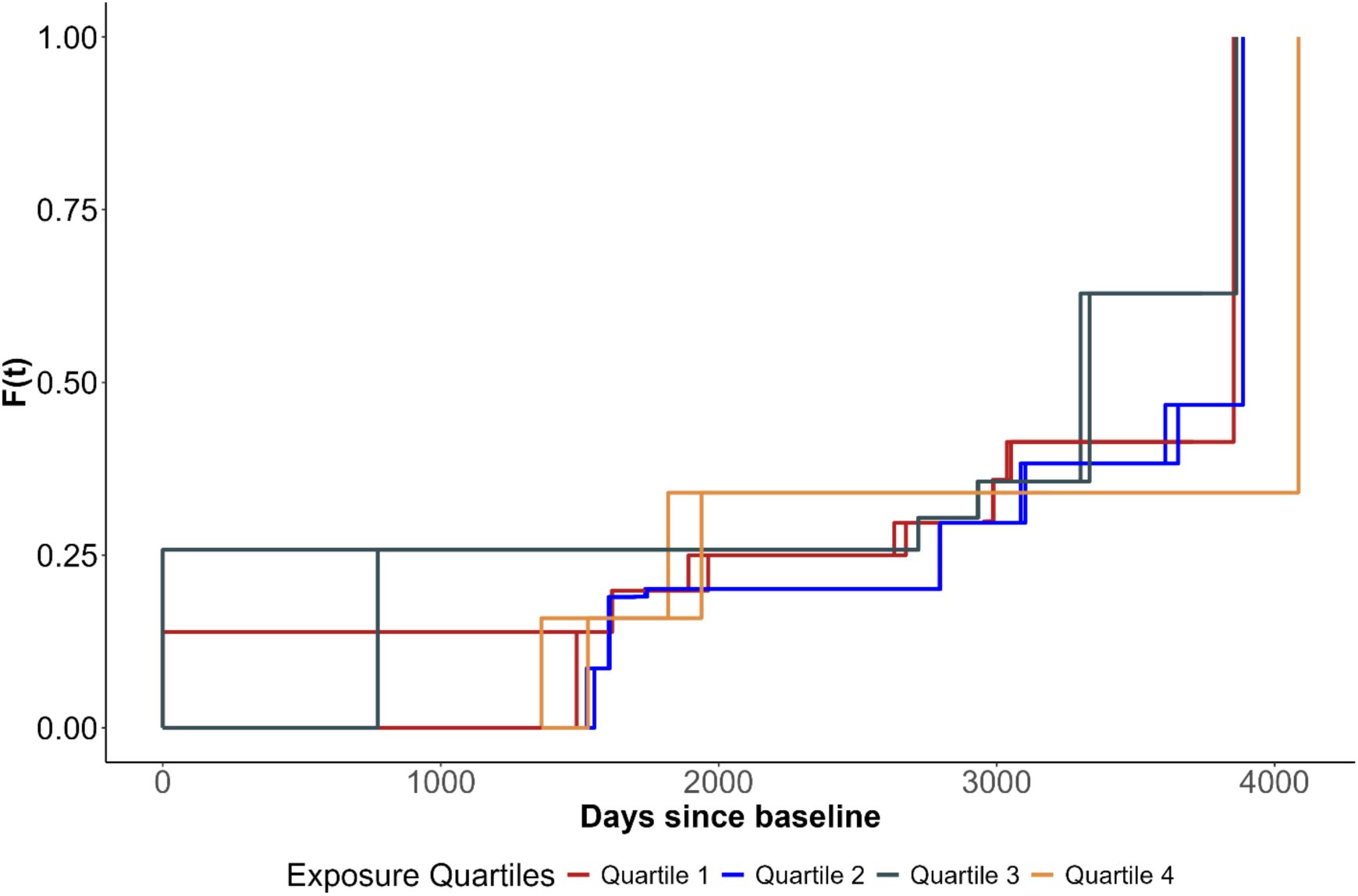
Cumulative incidence curves for incident aTRH by quartiles of urinary sodium. **Note:** The cumulative incidence curve shown above is the result from fitting a non-parametric maximum likelihood estimator (NPMLE) for univariate interval censored data guided by Turnbull’s estimator.

**Table 2.** Incident rates and hazard ratios of incident apparent treatment resistant hypertension for JHS participants with hypertension enrolled in 2000–2004, by quartiles of urinary sodium.

| Incident aTRH | Quartiles of urinary sodium |  |  |  | P Value for Trend |
| --- | --- | --- | --- | --- | --- |
|  | Quartile 1<br>(253 to 2530<br>mg)<br>N=113 | Quartile 2<br>(2553 to 3657<br>mg)<br>N=113 | Quartile 3<br>(3680 to 4692<br>mg)<br>N=113 | Quartile 4<br>(4715 to 9775<br>mg)<br>N=113 |  |
| Cases, n (%) | 29 (25.7%) | 28 (24.8%) | 33 (29.2%) | 33 (29.2%) | 0.166 |
| Event Summary,<br>By exam 2<br>(cases/no. at risk) | 17/96 | 16/100 | 25/97 | 19/102 |  |
| By exam 3<br>(cases/no. at risk) | 12/64 | 12/75 | 8/64 | 14/73 |  |
| HR (95% CI) |  |  |  |  |  |
| Model 1† | 1.00 (reference) | 0.88<br>(0.52, 1.51) | 1.14<br>(0.67, 1.94) | 1.04<br>(0.61, 1.80) |  |
| Model 2§ | 1.00 (reference) | 0.66<br>(0.32, 1.35) | 0.95<br>(0.50, 1.82) | 0.87<br>(0.44, 1.70) |  |
| Model 3\|\| | 1.00 (reference) | 0.71<br>(0.34, 1.46) | 1.02<br>(0.50, 2.06) | 0.95<br>(0.46, 2.00) |  |

n (%) represents count (percentage)
HR=Hazard ratio. CI: confidence interval.
Standard errors were estimated using
† Includes adjustment for age and sex
§ Includes variables in model 1 and additional adjustment for smoking status, alcohol consumption, physical activity, BMI, diabetes, history of coronary heart disease, history of stroke, heart failure, chronic kidney disease, estimated glomerular filtration rate, low- and high-density lipoproteins, urinary albumin-to-creatinine ratio, medication adherence, and an indicator for each of six broad classes of antihypertensive medication used (ACEI, ARB, beta-blockers, calcium channel blockers, diuretics, and others).
|| Includes variables in models 1 and 2 and additional adjustment for income, education, and insurance status.

In a fully adjusted model (Model 3), the associations between 24-h urinary sodium excretion and incident aTRH did not reach statistical significance. Relative to Q1, the HRs with 95% confidence intervals (CIs) for Q2, Q3 and Q4 were 0.71 (0.34, 1.46), 1.02 (0.50, 2.06), and 0.95 (0.46, 2.00), respectively (P=0.166). As shown in **Figure 3**, the HR for incident aTRH across restricted quadratic splines of urinary sodium (mg/day) indicated a non-linear association. While both very low and very high 24-h urinary sodium excretion levels were associated with a lower risk of aTRH, the highest risk occurred within the Q3 range.

**Figure 3.**
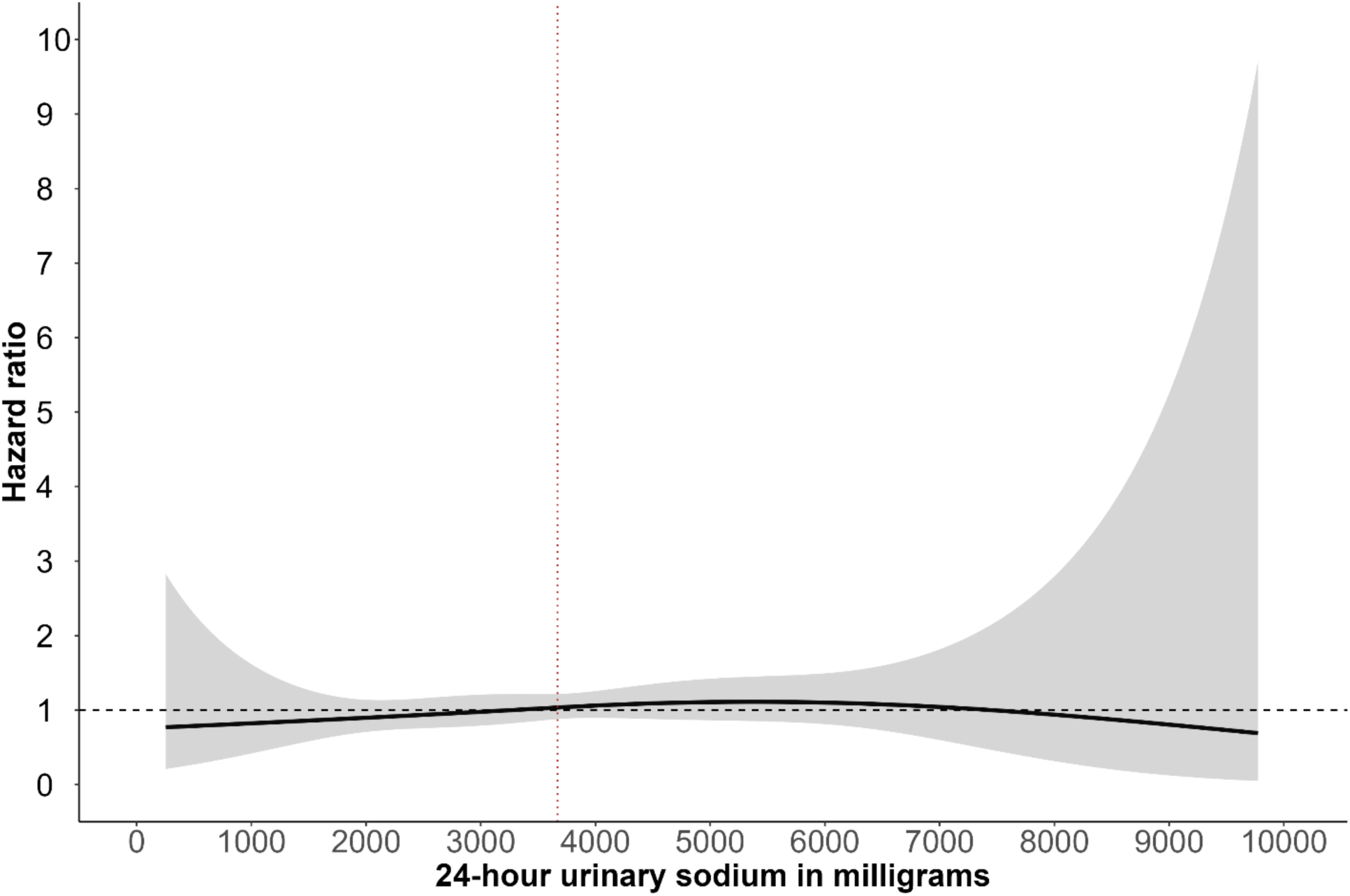
Hazard ratio for incident aTRH associated with urinary sodium by using restricted quadratic splines. Note: The hazard ratio for incident aTRH across restricted quadratic splines of urinary sodium (mg) with a single knot specified at the median value (red vertical line). The fitted model was a semi-parametric univariate proportional hazards model suited for interval censored data.

## DISCUSSION

This study examined the association between urinary sodium and incident aTRH among African American adults. Over a median follow-up of ∼7.5 years, 123 participants (27.2%) developed aTRH and there was no statistically significant association between urinary sodium and incident aTRH. The median 24-h urinary sodium excretion among JHS participants with hypertension was 3,669 mg/day, exceeding both the AHA recommendation of <2300 mg/day, the national average intake of 3,400mg/day^7^, and other similar cohort studies.^34–36^ This finding reinforces evidence of high sodium consumption among AA adults,^37^ with critical implications for the increasing incidence of hypertension-related complications, such as stroke and heart disease, in this population.^38^ This finding also underscores the urgent need for targeted public health interventions to reduce sodium intake within the AA community to prevent worsening health disparities.

A fully adjusted model did not show a statistically significant association between 24-hour urinary sodium excretion and incident aTRH over a median follow-up of ∼7.5 years. The HRs for Q2, Q3, and Q4 compared to Q1 did not show a significant increase in incident aTRH risk, suggesting that other factors may mediate the relationship between sodium intake and incident aTRH. The lack of association between urinary sodium and aTRH suggests that the role of sodium in hypertension-related outcomes may be more complex than previously thought^35^ and may involve other factors such as genetics, age, and dietary components like potassium.^39^ Therefore, a comprehensive approach to hypertension management, one that considers multiple factors beyond sodium intake, is needed to effectively address the risk of aTRH.

Baseline characteristics revealed statistically significant differences between participants by sodium levels, highlighting the complex interplay between sodium intake and demographic, lifestyle and clinical factors. Participants with higher sodium levels were younger, more likely female, had higher income, consumed alcohol, adhered to antihypertensive medications, had higher BMI, higher eGFR, and lower HDL cholesterol. Conversely, higher sodium levels were less prevalent among participants with history of stroke, chronic kidney disease, or those using ACE inhibitors or beta-blockers. These findings suggest an association between higher sodium intake and favorable socioeconomic status and certain health behaviors^40^, but also with adverse metabolic profiles.^41,42^ This duality of association, where high sodium is linked to both favorable lifestyle behaviors and adverse health outcomes, has been shown in other studies.^39^

The primary strength of this study is that it is a prospective, community-based observational study examining the association between urinary sodium and incident aTRH among AA adults in the United States. By using a biological measure of sodium, this study offers a more reliable assessment compared to self-reported dietary recall methods. However, some limitations should be acknowledged. First, the relatively small sample size and the limited number of aTRH cases may have reduced the statistical power to detect statistically significant associations. Second, the observational nature of the study precludes causal inferences, as residual confounding by unmeasured variables cannot be ruled out.^43^ Third, many participants had missing urinary sodium data, and sodium excretion was measured at a single 24-hour period, which may not accurately reflect long-term intake,^44,45^ as repeated 24-hour urine collections were not used in this study. Finally, there were only 2 follow-up measurements, and this may not accurately reflect the true incidence of aTRH.

This study has important research and policy implications. Future research should employ longitudinal designs with repeated 24-h urine collection to better capture measures of long-term sodium intake on BP. Larger studies are also needed to provide more robust evidence of the association between sodium intake and aTRH, as the small sample size and limited number of aTRH cases in our analysis may have limited the statistical power to detect significant associations. Further, multiple factors associated with hypertension including genetic, lifestyle and dietary components, should be considered, with large randomized controlled trials to establish causality. Subgroup analyses could tailor public health recommendations to specific populations, and identifying biomarkers that associate sodium to BP will help in developing targeted interventions. Finally, evaluating the effectiveness of public health strategies aimed at reducing sodium intake and their associations with hypertension prevalence and outcomes will be essential for informing more effective prevention and treatment strategies.

## PERSPECTIVES

This community-based cohort study highlights the high sodium intake among AA adults with hypertension. While the association between 24-hour urinary sodium excretion and incident aTRH did not reach statistical significance, the findings highlight the need for targeted public health interventions to reduce sodium consumption in the AA populations. Larger, longitudinal studies are needed to confirm these findings and explore the complex associations between sodium intake, other dietary components, lifestyle factors, and genetics in hypertension management. Addressing these gaps could help reduce health disparities and improve health outcomes for AA adults.

## Data Availability

The authors do not have permission to share data.

https://www.jacksonheartstudy.org/Research/Study-Data/Data-Access

## Acknowledgement

The authors wish to thank the staff and participants of the Jackson Heart Study (JHS).

## Sources of Funding

The Jackson Heart Study is supported and conducted in collaboration with Jackson State University (N01-HC-95170), University of Mississippi Medical Center (N01-HC-95171), and Touglaoo College (N01-HC-95172) and contracts HHSN268201300046C, HHSN268201300047C, HHSN268201300048C, HHSN268201300049C, and HHSN268201300050C from the National Heart, Lung, and Blood Institute and the National Institute on Minority Health and Health Disparities at the National Institutes of Health. This work was also supported by the National Institutes of Health (HL047540, HL117323, HL117323-02S2, K24-HL125704) from the National Heart, Lung, and Blood Institute, 5KL2TR001065 from the National Center for Advancing Translational Sciences, Bethesda, MD, and 15SFRN2390002 from the American Heart Association. The funding sources played no role in the design, conduct, analyses, or reporting of the present study.

## Disclosures

None

## Notes

### Competing Interest Statement

The authors have declared no competing interest.

### Author Declarations

The study was approved by the Institutional Review Boards of Jackson State University, Tougaloo College, and the University of Mississippi Medical Center in Jackson. All participants provided written informed consent. This article was also reviewed by the JHS Publications and Presentations Committee and has been approved for publication.

## References

1. Chobufo MD, Gayam V, Soluny J, Rahman EU, Enoru S, Foryoung JB, Agbor VN, Dufresne A, Nfor T. Prevalence and control rates of hypertension in the USA: 2017–2018. International Journal of Cardiology: Hypertension. 2020;6:100044–100044. doi: 10.1016/j.ijchy.2020.100044

2. Mozaffarian D, Benjamin E, Go A, Arnett D, Blaha M, Cushman M, de Ferranti S. Heart disease and stroke statistics—2015 update: a report from the American Heart Association. Circulation. 2015;131:e29–e322.

3. Kochanek KD, Arias E, Anderson RN. How Did Cause of Death Contribute to Racial Differences in Life Expectancy in the United States in 2010? Key findings. NCHS Data Brief. 2013:1–8.

4. Appel L. Another major role for dietary sodium reduction: improving blood pressure control in patients with resistant hypertension. Hypertension. 2009;54:444–446.

5. Pimenta E, Gaddam K, Oparil S, Aban I, Husain S, Dell’Italia L, Calhoun D. Effects of dietary sodium reduction on blood pressure in subjects with resistant hypertension: results from a randomized trial. Hypertension. 2009;54:475–481.

6. Aburto N, Ziolkovska A, Hooper L, Elliot P, Cappuccio F, Meerpohl J. Effect of lower sodium intake on health: Systematic review and meta-analysis. BMJ. 2013;346:f1326.

7. Cogswell M, Loria C, Terry A, Zhao L, Wang C, Chen T, Wright J, Pfeiffer C, M’erritt R, Moy C, et al. Estimated 24-hour urinary sodium and potassium excretion in US adults. JAMA. 2018;319:1209–1220.

8. Jackson S, King S, Zhao L, Cogswell M. Prevalence of excess sodium intake in the United States—NHANES, 2009–2012. Morb Mortal Wkly Rep. 2016;64:1393–1397.

9. Airhihenbuwa C, Kumanyika S, Agurs T, Lowe A, Saunders D, Morssink C. Cultural aspects of African American eating patterns. Ethnicity & Health. 1996;1:245–260.

10. Adinkrah E, Bazargan M, Wisseh C, Assari S. Adherence to Hypertension Medications and Lifestyle Recommendations among Underserved African American Middle-Aged and Older Adults. Int J Environ Res Public Health. 2020;17:6538. doi: 10.3390/ijerph17186538

11. Wielgosz A, Robinson C, Mao Y, Jiang Y, Campbell N, Muthuri S, Morrison H. The impact of using different methods to assess completeness of 24-hour urine collection on estimating dietary sodium. The Journal of Clinical Hypertension. 2016;18:581–584.

12. Lucko A, Doktorchik C, Woodward M, Cogswell M, Neal B, Rabi D, Anderson C, He F, MacGregor G, L’Abbe M, et al. Percentage of ingested sodium excreted in 24-hour urine collections: a systematic review and meta-analysis. The Journal of Clinical Hypertension. 2018;20:1220–1229.

13. Calhoun D, Jones D, Textor S, Goff D, Murphy T, Toto R, White A, Cushman W, White W, Sica D, et al. Resistant hypertension: diagnosis, evaluation, and treatment: a scientific statement from the American Heart Association Professional Education Committee of the Council for High Blood Pressure Research. Hypertension. 2008;51:1403– 1419.

14. Langford A, Akinyelure O, M’oore Jr T, Howard G, Min Y, Hillegass W, Bress A, Tajeu G. Under-utilization of treatment for blacks with apparent treatment-resistant hypertension: Jackson Heart Study and the REasons for Geographic And Racial Differences in Stroke Study. Hypertension. 2020;76:1600–1607.

15. Spence J, Rayner B. Hypertension in blacks: individualized therapy based on renin/aldosterone phenotyping. Hypertension. 2018;72:263–269.

16. Carpenter M, Crow R, Steffes M, Rock W, Heilbraun J, Evans G, Skelton T, Jensen RW, Sarpong D. Laboratory, reading center, and coordinating center data management methods in the Jackson Heart Study. Am J Med Sci 2004;328:131–144.

17. Fuqua S, Wyatt S, Andrew M, Sarpong D, Cunningham M, Taylor H. Recruiting African-American research participation in the Jackson heart study: methods, response rates, and sample description. Ethn Dis. 2005;15:S6–18.

18. Taylor H, Wilson J, Jones D, Sarpong D, Srinivasan A, Garrison R, Nelson C, Wyatt S. Toward resolution of cardiovascular health disparities in African Americans: design and methods of the Jackson heart study. Ethn Dis. 2005;15:S6–4.

19. Dubbert PM, Carithers T, Ainsworth BE, Taylor HA Jr, Wilson G, SB. W. Physical activity assessment methods in the Jackson Heart Study. Ethn Dis. 2005;15:S6–61.

20. Lloyd-Jones D, Hong Y, Labarthe D, Mozaffarian D, Appel L, Horn L, Greenlund K, Daniels S, Nichol G, Tomaselli G. Defining and setting national goals for cardiovascular health promotion and disease reduction: the American Heart Association’s strategic impact goal through 2020 and beyond. Circulation. 2010;121:586–613. doi: 10.1161/CIRCULATIONAHA.109.192703

21. Seals S, Colantonio L, Tingle J, Shimbo D, Correa A, Griswold M, Muntner P. Calibration of blood pressure measurements in the Jackson Heart Study. Blood Press Monit. 2019;24:130–136. doi: 10.1097/MBP.0000000000000379

22. Tanner R, Shimbo D, Irvin M, Spruill T, Bromfield S, Seals S, Young B, Muntner P. Chronic kidney disease and incident apparent treatment-resistant hypertension among blacks: Data from the Jackson Heart Study. The Journal of Clinical Hypertension. 2017;19:1117–1124.

23. Wang W, Young B, Fulop T, de Boer I, Boulware L, Katz R, Correa A, Griswold M. Effects of serum creatinine calibration on estimated renal function in African Americans: the Jackson Heart Study. Am J Med Sci 2015;349:379–384.

24. Levey A, Bosch J, Lewis J, Greene T, Rogers N, Roth D. A more accurate method to estimate glomerular filtration rate from serum creatinine: a new prediction equation. Modification of Diet in Renal Disease Study Group. Ann Intern Med. 1999;130:461–470. doi: 10.7326/0003-4819-130-6-199903160-00002

25. Sims K, Sims M, Glover L, Smit E, Odden M. Perceived discrimination and trajectories of C-reactive protein: The Jackson Heart Study. American Journal of Preventive Medicine. 2020;58:199–207.

26. Austin P, White I, Lee D, van Buuren S. Missing Data in Clinical Research: A Tutorial on Multiple Imputation. Statistics in Medicine. 2021;40:4198–4219.

27. White I, Royston P. Imputing missing covariate values for the Cox model. Statistics in Medicine. 2009;28:1982–1998.

28. Rubin D. Multiple imputation for nonresponse in surveys. New York, USA: John Wiley & Sons; 2004.

29. Finkelstein D. A proportional hazards model for interval-censored failure time data. Biometrics. 1986;42:845–854.

30. Turnbull B. The empirical distribution function with arbitrarily grouped, censored and truncated data. Journal of the Royal Statistical Society Series B (Methodological). 1976;38:290–300.

31. R Core Team. A Language and Environment for Statistical Computing. R Foundation for Statistical Computing. https://www.R-project.org. 2023.

32. Anderson-Bergman C. icenReg: regression models for interval censored data in R. Journal of Statistical Software 2017;81:1–23.

33. Buuren S, Groothuis-Oudshoorn K. mice: Multivariate Imputation by Chained Equations in R. Journal of Statistical Software. 2011;45:1–67.

34. Elfassy T, Chamany S, Bartley K, Yi S, Angell S. Lower 24-h urinary sodium excretion is associated with hypertension control: the 2010 Heart Follow-Up Study. Journal of Human Hypertension. 2020;34:624–632.

35. Elliott P, Muller D, Schneider-Luftman D, Pazoki R, Evangelou E, Dehghan A, Neal B, Tzoulaki I. Estimated 24-hour urinary sodium excretion and incident cardiovascular disease and mortality among 398 628 individuals in UK Biobank. Hypertension. 2020;76:683–691.

36. Jarrar A, Stojanovska L, Apostolopoulos V, Cheikh Ismail L, Feehan J, Ohuma E, Ahmad A, Alnoaimi A, Al Khaili L, Allowch N, et al. Assessment of sodium knowledge and urinary sodium excretion among regions of the United Arab Emirates: a cross-sectional study. Nutrients. 2020;12:2747.

37. Fulgoni V, Agarwal S, Spence L, Samuel P. Sodium intake in US ethnic subgroups and potential impact of a new sodium reduction technology: NHANES Dietary Modeling. Nutrition Journal. 2014;13:1–9.

38. Lloyd-Jones D, Adams R, Brown T, Carnethon M, Dai S, De Simone G, Ferguson T, Ford E. American Heart Association Statistics Committee and Stroke Statistics Subcommittee. Heart disease and stroke statistics--2010 update: a report from the American Heart Association. Circulation. 2010;121:e46–e215.

39. Mente A, O’Donnell M, Rangarajan S, McQueen M, Poirier P, Wielgosz A, Morrison H, Li W, Wang X, Di C, et al. Association of urinary sodium and potassium excretion with blood pressure. New England Journal of Medicine. 2014;371:601–611.

40. Nisa H, Kurotani K. Diet quality, socioeconomic differences, and health disparities. Frontiers in Nutrition. 2023;10:1250439.

41. Baudrand R, Campino C, Carvajal C, Olivieri O, Guidi G, Faccini G. High sodium intake is associated with increased glucocorticoid production, insulin resistance and metabolic syndrome. Clinical Endocrinology. 2014;80:677–684.

42. Oh S, Han K, Han S, Koo H, Kim S, Chin H. Association of sodium excretion with metabolic syndrome, insulin resistance, and body fat. Medicine,. 2015;94:e1650.

43. Anderson C, Delker E, Ix J. Sodium and health outcomes: Ascertaining valid estimates in research studies. Current Atherosclerosis Reports. 2021;23:35.

44. Mente A, O’Donnell M, Rangarajan S, Dagenias G, Lear S, M’cQueen M. Associations of urinary sodium excretion with cardiovascular events in individuals with and without hypertension: A pooled analysis of data from four studies. The Lancet. 2016;388 465–475.

45. He FJ, MacGregor G. Reducing population salt intake worldwide: From evidence to implementation. Progress in Cardiovascular Diseases. 2018;61:94–98.

